# Efficacy and Safety of Edoxaban in Anticoagulant Therapy Early after Surgical Bioprosthetic Valve Replacement: the ENBALV trial

**DOI:** 10.1101/2024.10.16.24315630

**Authors:** Chisato IzumI, Masashi Amano, Satsuki Fukushima, Hitoshi Yaku, Kiyoyuki Eishi, Taichi Sakaguchi, Nobuhisa Ohno, Arudo Hiraoka, Kenji Okada, Yoshikatsu Saiki, Takashi Miura, Tatsuhiko Komiya, Manabu Minami, Haruko Yamamoto, Katsuhiro Omae, ENBALV Trial investigators

**Author notes:** A full list of the investigators in the ENBALV trial is provided in the Supplemental material. Address for correspondence: Chisato Izumi, MD, PhD, Director, Department of Heart Failure and Transplantation, National Cerebral and Cardiovascular Center, 6-1 Kishibe Shimmachi, Suita, Osaka 564-8565 Japan.

## Abstract

**Background:** Anticoagulant therapy with vitamin K antagonists is recommended in the current guidelines for 3 to 6 months following bioprosthetic valve replacement to prevent thromboembolic events, including in patients with sinus rhythm. However, in the era of direct oral anticoagulants (DOACs), there is a paucity of evidence regarding the efficacy and safety of DOACs in this patient group.

**Methods:** The ENBALV trial was an investigator-initiated, phase 3, randomized, open-label, multicenter study that aimed to evaluate the efficacy and safety of edoxaban compared to warfarin within 3 months following bioprosthetic valve replacement at the aortic and/or mitral position. The primary outcome was stroke or systemic embolism. The secondary outcomes included major bleeding, intracardiac thrombus, and a composite of stroke, systemic embolism, or major bleeding.

**Results:** Of 410 enrolled patients, 389 were included in the final analysis (73±6 years, 56.8% male, 79.4% sinus rhythm, edoxaban group: n=195, warfarin group: n=194). The primary outcome occurred in 0.5% (n=1) in the edoxaban group, whereas in 1.5% (n=3) in the warfarin group (risk difference, −1.03, 95% confidence interval [CI], −4.34 to 1.95). Major bleeding occurred in 4.1% (n=8) in the edoxaban group and in 1.0% (n=2) in the warfarin (risk difference, 3.07; 95% CI, −0.67 to 7.27). The incidence of major bleeding was numerically higher in the edoxaban group, but no fatal bleeding or intracranial hemorrhage was observed in patients treated with edoxaban, whereas one fatal intracranial hemorrhage occurred in the warfarin group. Intracardiac thrombus did not occur in any of the patients in the edoxaban group, but did occur in 1.0% (n=2) in the warfarin group.

**Conclusions:** Edoxaban had comparable efficacy to warfarin for the prevention of thromboembolic events in patients early after bioprosthetic valve replacement, suggesting that it is a potential alternative anticoagulant therapy.

**Trial registration:** The study was registered with the Japan Registry of Clinical Trials (jRCT), with reference number 2051210209 (30 March 2022; https://jrct.niph.go.jp/latest-detail/jRCT2051210209).

**Clinical perspective:** *What is new?:*

- Current guidelines recommend the administration of warfarin for patients early after bioprosthetic valve replacement, including those with sinus rhythm, but there is a paucity of evidence regarding the efficacy and safety of direct oral anticoagulants (DOACs) in this patient group.
- ENBALV trial provides the first large-scale evidence on the use of DOAC in patients early after bioprosthetic valve surgery.
- ENBALV trial demonstrated that edoxaban had comparable efficacy to warfarin for preventing thromboembolism. No fatal or intracranial hemorrhage was observed with edoxaban. Our results suggest that edoxaban could be an alternative to warfarin in this patient group.

*What are the clinical implications?:*

- The availability of edoxaban could offer more flexibility in anticoagulant treatment options after bioprosthetic valve surgery. Since edoxaban does not require regular and frequent blood testing, it could simplify the care process, reduce the burden on patients, and improve their quality of life, especially in the crucial early period after surgery.
- The availability of edoxaban also give benefits for medical stuffs as well as patients, because edoxaban can be used with constant dose, no need of routine monitoring of anticoagulation activity, and a low risk of interaction with other drugs and food.

## Introduction

The number of patients with valvular heart disease has been increasing and this trend has been paralleled by an increase in the prevalence of bioprosthetic valve replacement with aging society.^1^ The incidence of embolic events has been reported to be high early after bioprosthetic valve replacement;^2–6^ therefore, anticoagulant therapy with vitamin K antagonists is recommended in the current guidelines for 3 to 6 months following bioprosthetic valve replacement to prevent thromboembolic events, including in patients with sinus rhythm.^7–9^

Randomized clinical trials have demonstrated the efficacy and safety of direct oral anticoagulants (DOACs) versus warfarin for the treatment of patients with nonvalvular atrial fibrillation (AF),^10–12^ and DOACs have become widely used, because the routine monitoring of anticoagulation activity is not required and there is a low risk of interaction with other drugs and food.

Evidence supporting the use of DOACs in patients with AF who have undergone bioprosthetic valve replacement has been provided by several subgroup analyses of randomized clinical trials^13–15^ and observational studies^16–18^. In addition, a randomized clinical trial that compared the use of rivaroxaban and warfarin in patients with AF and a bioprosthetic mitral valve^19^ revealed the non-inferiority of rivaroxaban to warfarin with respect to a composite outcome of death, major cardiovascular events, or major bleeding. However, fewer than 20% (n=189) of the participants had undergone bioprosthetic valve replacement within the preceding 3 months, and there were no participants with sinus rhythm. In addition, another study demonstrated the efficacy and safety of edoxaban versus warfarin in patients who underwent mitral valve repair or bioprosthetic valve replacement, but the bioprosthetic valve replacement group was small (n=152).^20^

The efficacy and safety of EdoxabaN in anticoagulant therapy after surgical Bioprosthetic vALVe replacement (ENBALV) trial was a randomized clinical trial that aimed to evaluate the efficacy and safety of edoxaban compared to warfarin early after bioprosthetic valve replacement.

## Methods

### Trial design and oversight

The trial design has been described previously.^21^ Briefly, the ENBALV trial was an investigator-initiated, phase 3, randomized, open-label, multicenter study. We evaluated the efficacy and safety of the use of edoxaban compared to warfarin within 3 months following bioprosthetic valve replacement. Details of the participating investigators and trial organization are provided in the Supplementary material.

All the procedures involving human participants were performed in accordance with the 1964 Declaration of Helsinki and its later amendments or comparable ethical standards, and with the Japanese Pharmaceutical Affairs Act and related laws and regulations. The study protocol was approved by the National Cerebral and Cardiovascular Center Review Board in addition to the Institutional Review Board of each participating institution and it was registered with the Japan Registry of Clinical Trials (jRCT 2051210209). Written informed consent was obtained from all the patients before they were recruited.

An independent safety monitoring committee monitored all the safety data and was involved in decisions regarding trial continuation or protocol changes. All suspected outcomes and the results of imaging evaluations were adjudicated by an independent event committee, the members of which were unaware of the trial group assignments.

### Trial population

We studied patients aged 18 to 85 years who underwent bioprosthetic valve replacement at the aortic and/or mitral position. Patients with both sinus rhythm and AF were included. The main exclusion criteria were a contraindication to the use of either warfarin or edoxaban, an extremely high risk of hemorrhage, and the presence of a mechanical valve or greater than moderate mitral stenosis, except if replacement of the mechanical valve or stenotic mitral valve by a bioprosthetic valve was performed at this operation. Further details regarding the inclusion and exclusion criteria of the ENBALV trial are shown in Table S1. To minimize the risk of withdrawal because of worsening renal function after surgery, the patient selection criterion for renal function was set as a creatinine clearance of ≥30 mL/min, not ≥15 mL/min.

### Trial procedures

Eligible patients were randomly allocated to either the edoxaban or warfarin group at a 1:1 ratio using a web-based randomization system and the minimization method. The adjustment factors were 1) the valve position (aortic valve alone, mitral valve alone, or both valves), 2) the presence of AF, and 3) the administration of antiplatelet drugs.

Bioprosthetic valve replacement was performed within 8 weeks of randomization. Edoxaban or warfarin administration was started following bioprosthetic valve replacement, as soon as the surgeons had determined that it was appropriate for anticoagulant therapy to commence. Anticoagulant therapy initiation could be delayed according to patients’ condition such as postoperative wound bleeding. The present study was performed during the unstable period immediately following open heart surgery; therefore, the surgeons could use their discretion to determine when anticoagulant therapy should be initiated, to prioritize patient safety.

Edoxaban was orally administered at the dose of 60 mg once daily, or 30 mg once daily when the patient’s creatinine clearance was 30 to 50 mL/min (calculated using the Cockcroft–Gault equation), their body weight was ≤60 kg, or they were being concomitantly treated with a P-glycoprotein inhibitor. The dose of warfarin was adjusted under monitoring the prothrombin time-international normalized ratio (PT-INR). Administration of edoxaban or warfarin was continued for 12 weeks after surgery if the patient did not meet any of the criteria for its discontinuation, and their administration could be continued until 24 weeks at the surgeons’ discretion. Clinical evaluations were conducted from the initiation of study drug administration to the end of study treatment.

The patients underwent clinical assessment and laboratory testing 1 and 7 days after the administration of edoxaban or warfarin commenced and at discharge. After discharge, they were scheduled to visit an outpatient clinic 4, 8, and 12 weeks after initiation of anticoagulant therapy. Clinical assessments, including the evaluation of symptoms suggestive of clinical thromboembolic or hemorrhagic events and laboratory testing, were performed at the outpatient visits.

The patients underwent a 12-lead electrocardiography and transthoracic echocardiography as part of eligibility assessment prior to randomization, at discharge, and 12 weeks after initiation of anticoagulant therapy. Brain magnetic resonance imaging (MRI) or computed tomography (CT) was also performed during the eligibility assessment process prior to randomization and 12 weeks after initiation of anticoagulant therapy.

### Primary and secondary outcomes

The primary outcome was a composite of stroke or systemic embolism. The key secondary outcomes were major bleeding, intracardiac thrombus, and a composite of stroke, systemic embolism, or major bleeding, according to the definition of the International Society on Thrombosis and Haemostasis. The secondary outcomes also included the individual components of the composite outcome and other clinical events. A complete list of trial endpoints is provided in Table S2.

### Statistical analysis

The event rates for the primary endpoints in the study and control treatment groups were estimated to be 1% to 3%, based on historical reports.^2–5^ Thus, the difference in the event rate between the control and study treatment groups was not expected to be very large. Furthermore, the number of patients that could be included within a reasonable study period was estimated to be approximately 450, and it was judged not to be reasonable to conduct a non-inferiority trial on this scale to test the hypothesis. Therefore, to determine whether the study drug was not significantly inferior to the control drug, we decided to evaluate whether the difference in the point estimates of the event rate for the primary endpoint was below a certain threshold. A previous study^2^ that included data from a no-treatment group estimated that the event rate within 3 months after surgery in the absence of treatment was >7%. On this basis, the experts in our study group determined that 2% is a clinically acceptable and reasonable threshold for the difference in event rate between the treatment and control groups. Given the 450 enrolled patients, 1:1 allocation, and a dropout rate of approximately 10%, the primary endpoint was expected to be assessed in 202 patients in each group. The event rates *p* and *q* for the treatment and control groups were assumed to follow a uniform distribution within the interval [0.01, 0.03], and the occurrence of events in the treatment and control groups were assumed to follow a Bernoulli distribution with occurrence probabilities *p* and *q*, respectively. Under these conditions, the probability that the point estimate of *p−q* is <2% was evaluated by a simulation study and estimated to be approximately 90%. On the basis of the above considerations, the target number of patients to be enrolled in the trial was set as 450.

The primary analysis of the trial data was performed in a full analysis set (FAS) based on intention-to-treat principles. The FAS consisted of all the assigned study participants, excluding those who did not meet the primary enrollment criteria, those who never received any study treatments, those for whom post-assignment data were not available, and those who withdrew consent for the use of their data. The point estimate of the event rate for the primary endpoint was calculated for each of the study and control treatment groups to determine whether the difference in event rates was ≤2%. As an additional analysis, a similar evaluation was conducted on the per-protocol set, a population of the FAS that excludes subjects who were found to have violations or deviations from the study protocol that would affect the evaluation of the primary endpoint. Other endpoints were summarized and compared by groups, with subpopulation analyses conducted as necessary. All analyses were performed using SAS software, version 9.4 (SAS Institute Inc., Cary, NC, USA).

## Results

### Patient backgrounds and follow-up

Between May 6, 2022, and January 25, 2024, a total of 430 patients gave their consent to participate and were assessed for eligibility at 24 institutions. Of these, 410 patients underwent randomization (205 were assigned to receive edoxaban and 205 were assigned to receive warfarin). Of these 410 patients, 21 did not receive edoxaban or warfarin; therefore, 389 patients were included in the final analysis (edoxaban group, n=195; warfarin group, n=194) (Figure 1).

**Figure 1.**
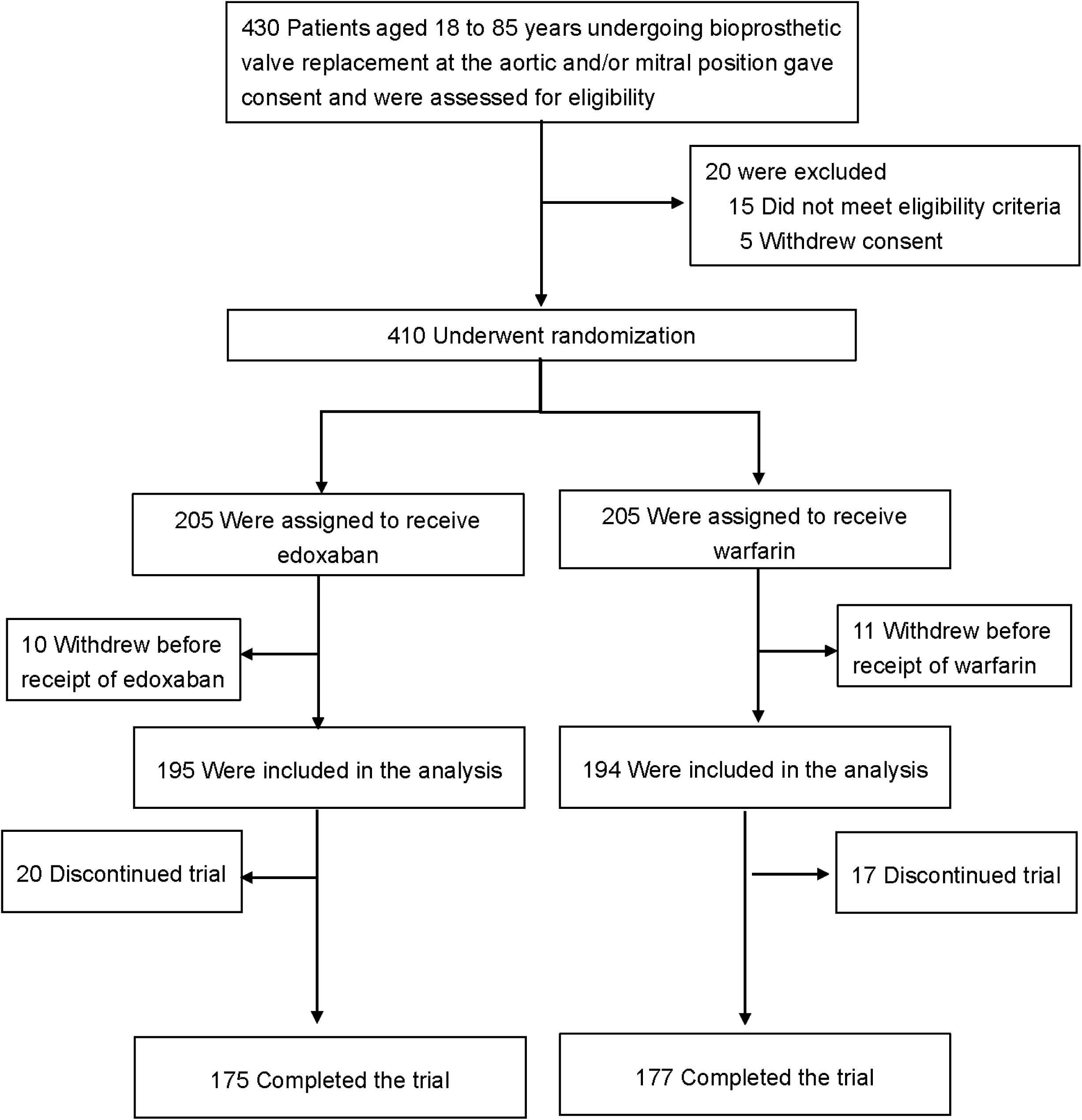
Enrollment, randomization, and follow-up of the patients.

The clinical characteristics of the patients are shown in Table 1. The two groups were well balanced with respect to their baseline characteristics. The mean age was 73 years, including 42 (10.8%) patients of ≥80 years of age, and 56.8% were male. The CHADS_2_ score, CHA_2_DS_2_-VASc score, and HAS-BLED score were 2.0±1.2, 3.4±1.4, and 1.1±0.6, respectively. Positions of bioprosthetic valve were aortic valve only in 339 (87.1%) patients, mitral valve only in 37 (9.5%) patients, and both in 13 (3.3%) patients. Of the 389 patients, 67 (17.2%) were taking antiplatelet drugs and 80 (20.6%) had atrial fibrillation, namely, the majority of patients had sinus rhythm.

**Table 1:**
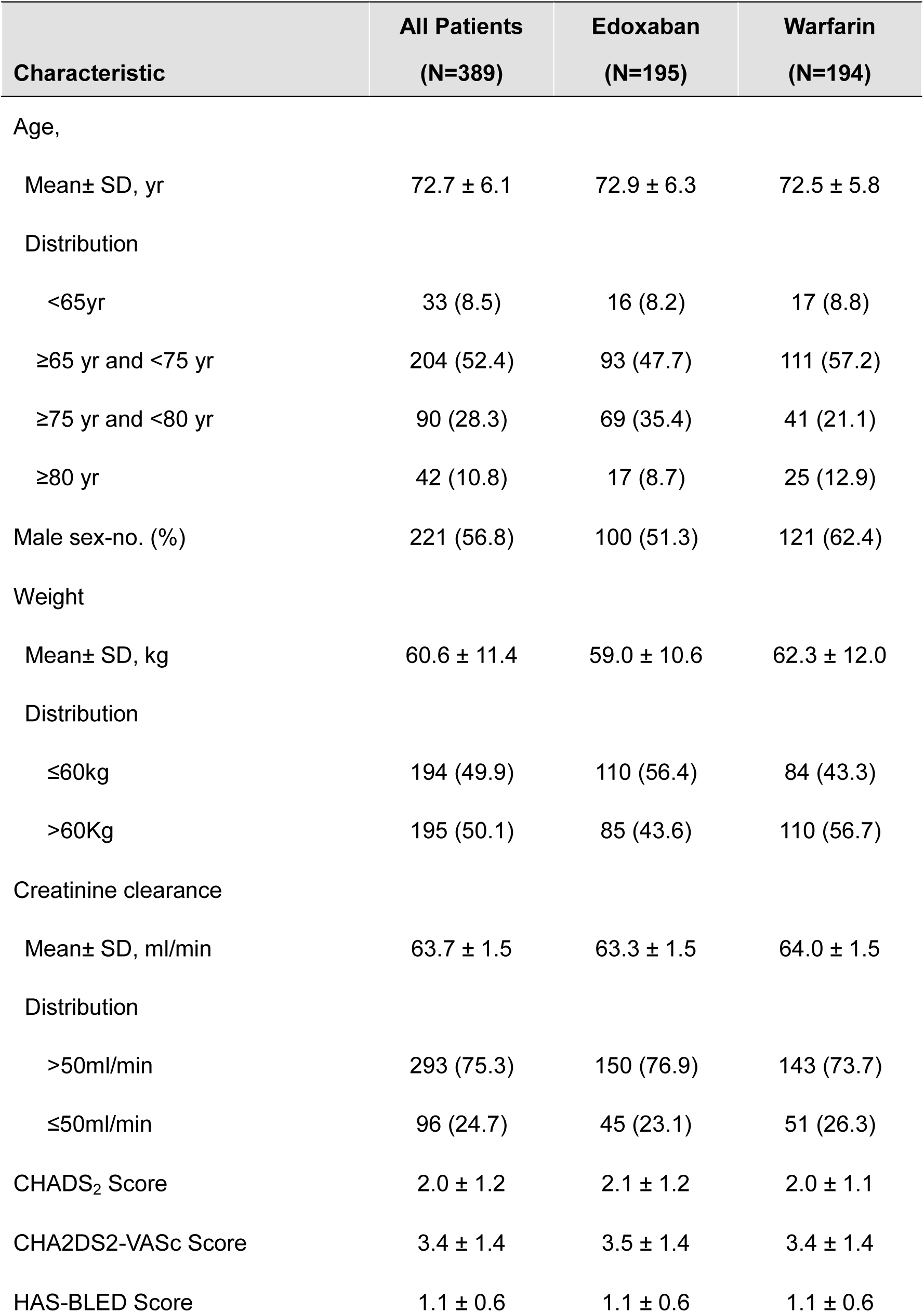
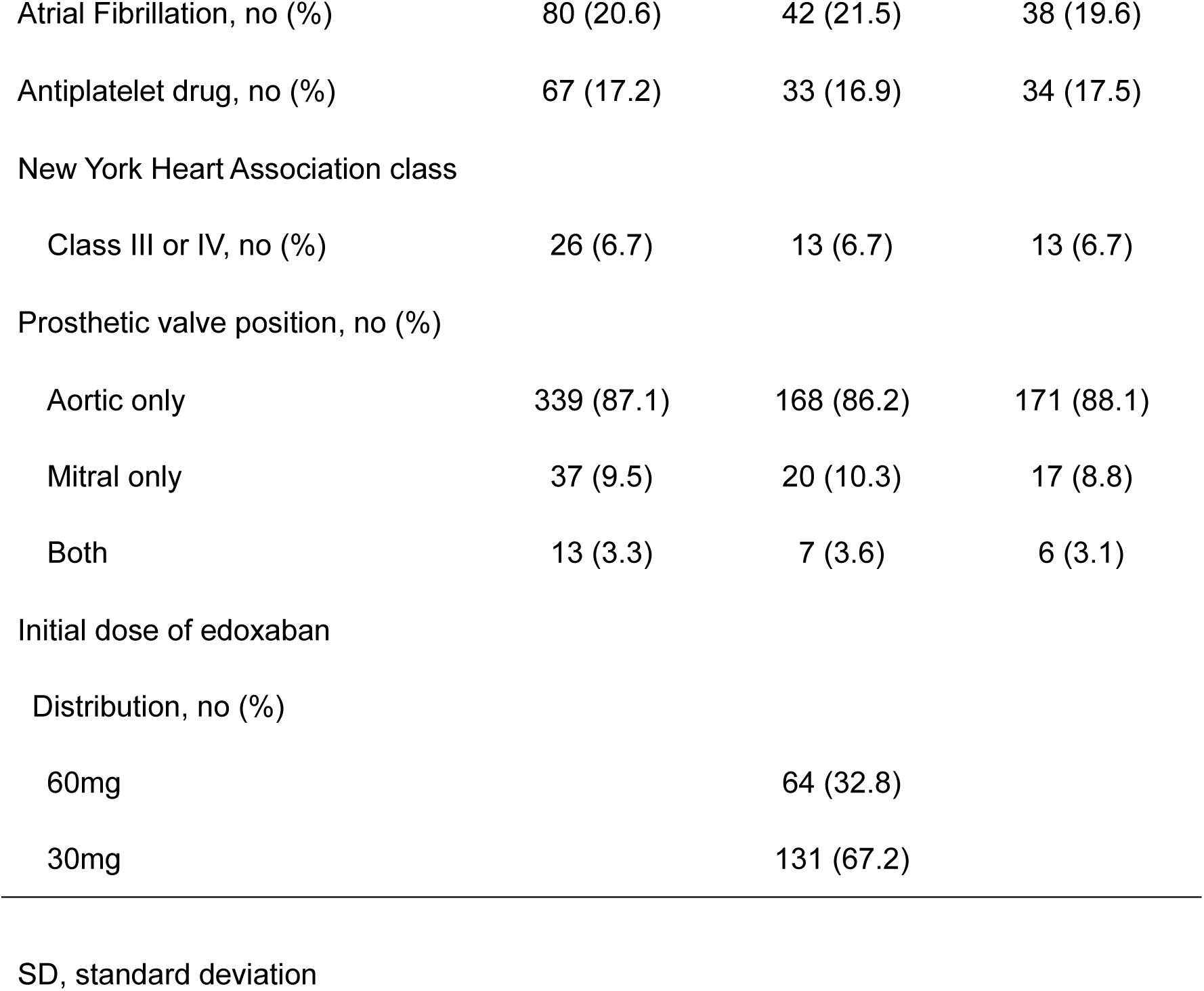
Demographic and Clinical Characteristics of the Patients.

| Characteristic | All Patients<br>(N=389) | Edoxaban<br>(N=195) | Warfarin<br>(N=194) |
| --- | --- | --- | --- |
| Age, |  |  |  |
| Mean± SD, yr | 72.7 ± 6.1 | 72.9 ± 6.3 | 72.5 ± 5.8 |
| Distribution |  |  |  |
| <65yr | 33 (8.5) | 16 (8.2) | 17 (8.8) |
| ≥65 yr and <75 yr | 204 (52.4) | 93 (47.7) | 111 (57.2) |
| ≥75 yr and <80 yr | 90 (28.3) | 69 (35.4) | 41 (21.1) |
| ≥80 yr | 42 (10.8) | 17 (8.7) | 25 (12.9) |
| Male sex-no. (%) | 221 (56.8) | 100 (51.3) | 121 (62.4) |
| Weight |  |  |  |
| Mean± SD, kg | 60.6 ± 11.4 | 59.0 ± 10.6 | 62.3 ± 12.0 |
| Distribution |  |  |  |
| ≤60kg | 194 (49.9) | 110 (56.4) | 84 (43.3) |
| >60Kg | 195 (50.1) | 85 (43.6) | 110 (56.7) |
| Creatinine clearance |  |  |  |
| Mean± SD, ml/min | 63.7 ± 1.5 | 63.3 ± 1.5 | 64.0 ± 1.5 |
| Distribution |  |  |  |
| >50ml/min | 293 (75.3) | 150 (76.9) | 143 (73.7) |
| ≤50ml/min | 96 (24.7) | 45 (23.1) | 51 (26.3) |
| CHADS<sub>2</sub> Score | 2.0 ± 1.2 | 2.1 ± 1.2 | 2.0 ± 1.1 |
| CHA<sub>2</sub>DS<sub>2</sub>-VASc Score | 3.4 ± 1.4 | 3.5 ± 1.4 | 3.4 ± 1.4 |
| HAS-BLED Score | 1.1 ± 0.6 | 1.1 ± 0.6 | 1.1 ± 0.6 |
| Atrial Fibrillation, no (%) | 80 (20.6) | 42 (21.5) | 38 (19.6) |
| Antiplatelet drug, no (%) | 67 (17.2) | 33 (16.9) | 34 (17.5) |
| New York Heart Association class |  |  |  |
| Class III or IV, no (%) | 26 (6.7) | 13 (6.7) | 13 (6.7) |
| Prosthetic valve position, no (%) |  |  |  |
| Aortic only | 339 (87.1) | 168 (86.2) | 171 (88.1) |
| Mitral only | 37 (9.5) | 20 (10.3) | 17 (8.8) |
| Both | 13 (3.3) | 7 (3.6) | 6 (3.1) |
| Initial dose of edoxaban |  |  |  |
| Distribution, no (%) |  |  |  |
| 60mg |  | 64 (32.8) |  |
| 30mg |  | 131 (67.2) |  |

SD, standard deviation

The date of the last patient follow-up was April 25, 2024. The median duration of participation in the trial was 98 days (interquartile range, 92 to 106 days). Of the 389 patients, 20 in the edoxaban group (11 discontinued edoxaban because of adverse events, and nine were judged as lack of capability to continue participation in the trial) and 17 in the warfarin group (six discontinued warfarin because of adverse events, two withdrew their consent, eight were judged as lack of capability to continue participation in the trial, and one died) did not complete the trial.

The patients in the warfarin group had PT-INR values within the therapeutic range (2.0–3.0 seconds) for a median of 19.0% (interquartile range, 7.0%–31.4%) of the study period.

### Primary and secondary outcomes

The primary outcome occurred in one patient (0.5%) in the edoxaban group, whereas in three patients (1.5%) in the warfarin group (risk difference, −1.03; 95% confidence interval [CI], −4.34 to 1.95) (Figures 2 and 3A). Systemic embolism did not occur in any of the study patients; thus all of the events of primary outcome were stroke. Intracardiac thrombus did not occur in any of the patients in the edoxaban group, but did occur in two patients (1.0%) in the warfarin group (risk difference, −1.03; 95% CI, −4.07 to 1.52) (Figures 2 and 3B).

**Figure 2.**
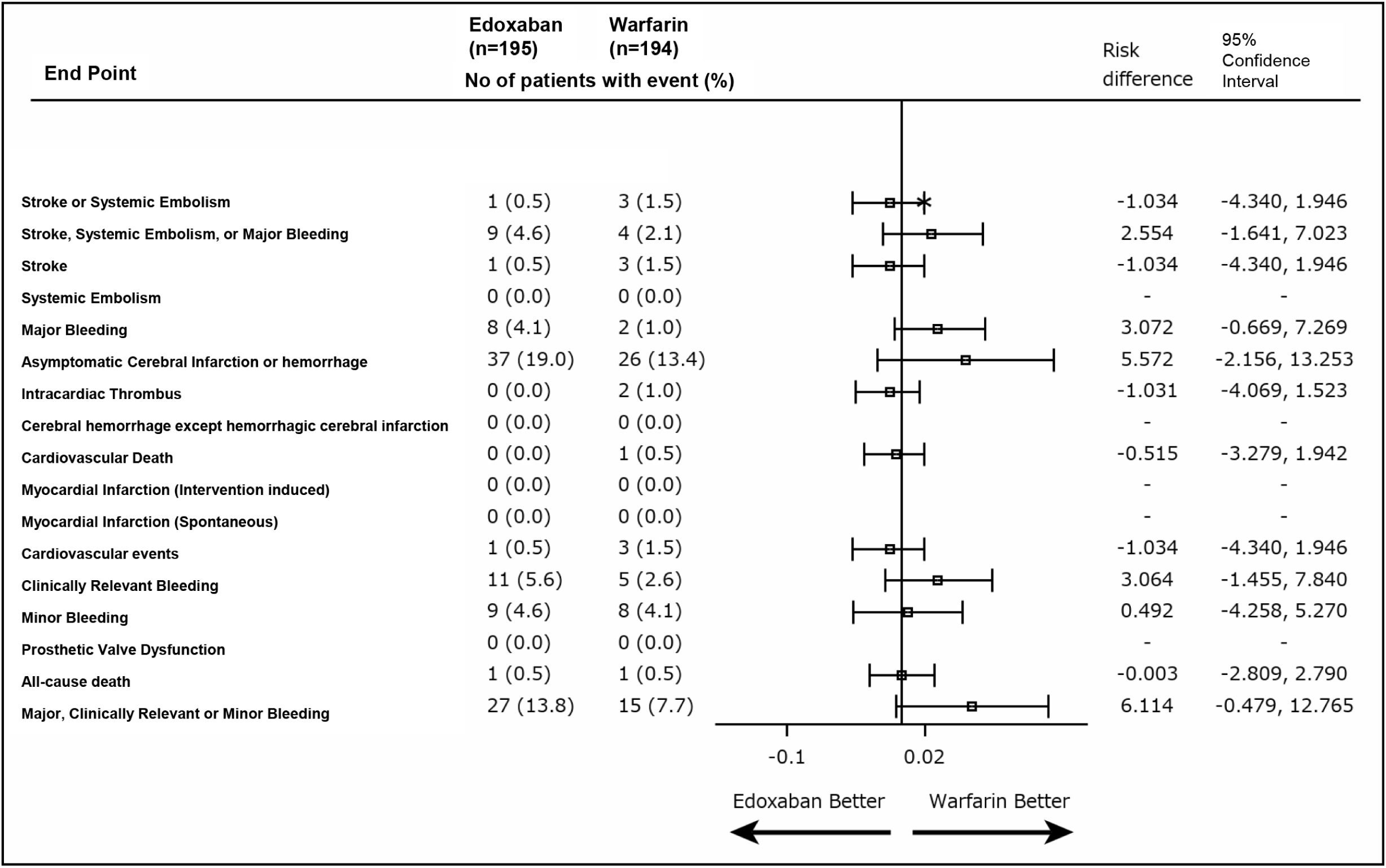
Primary and secondary endpoints.

**Figure 3.**
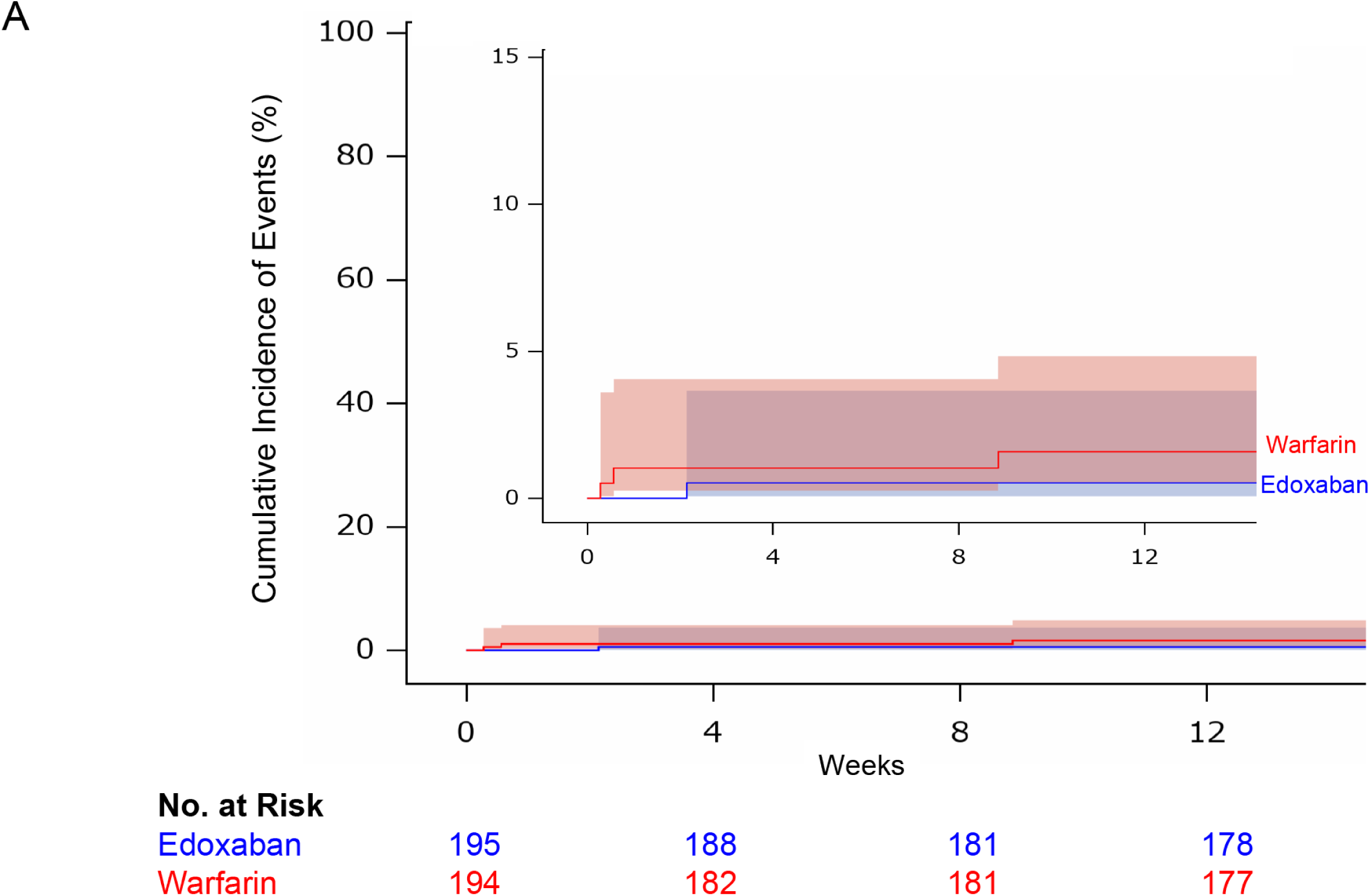
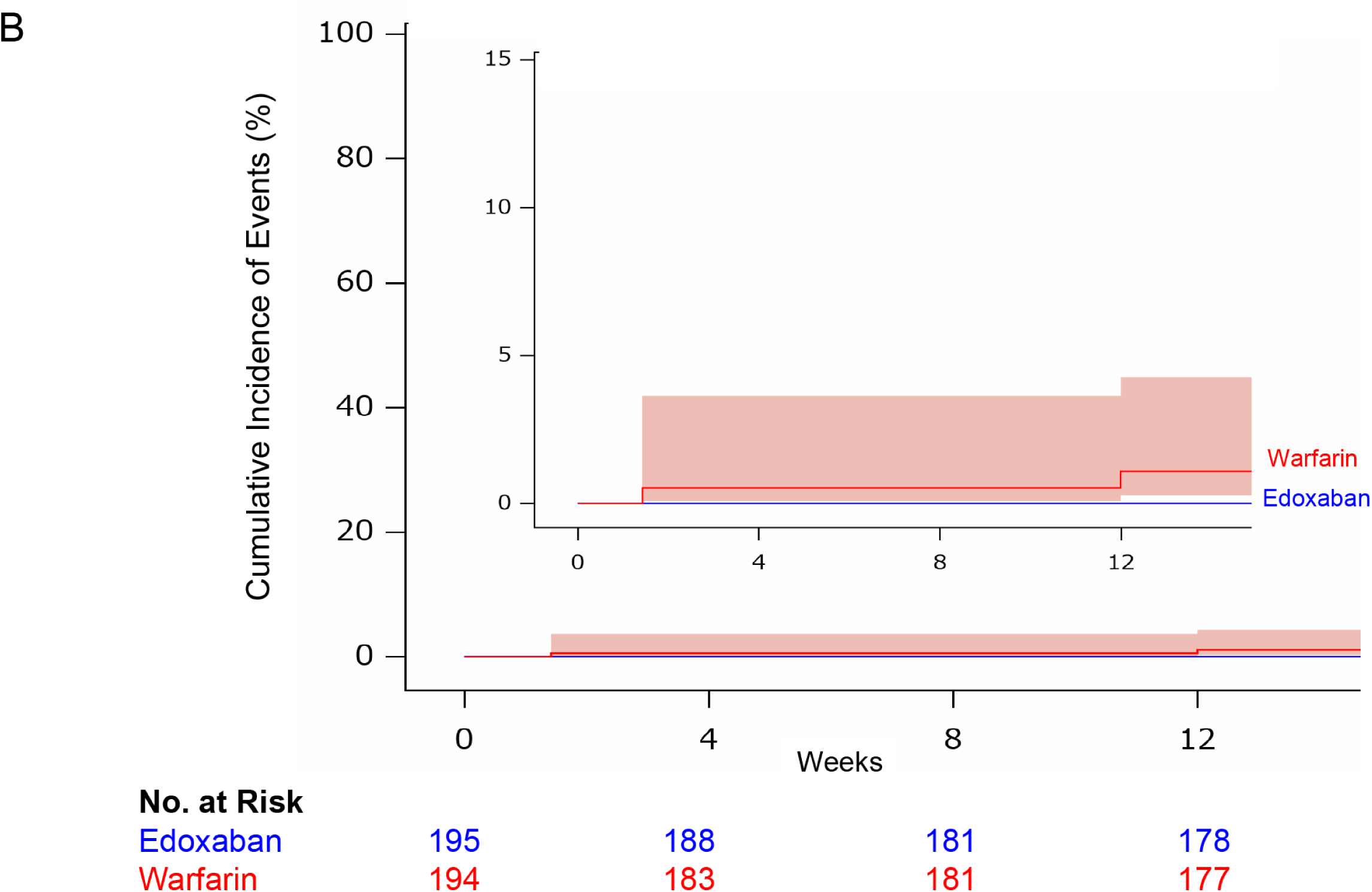
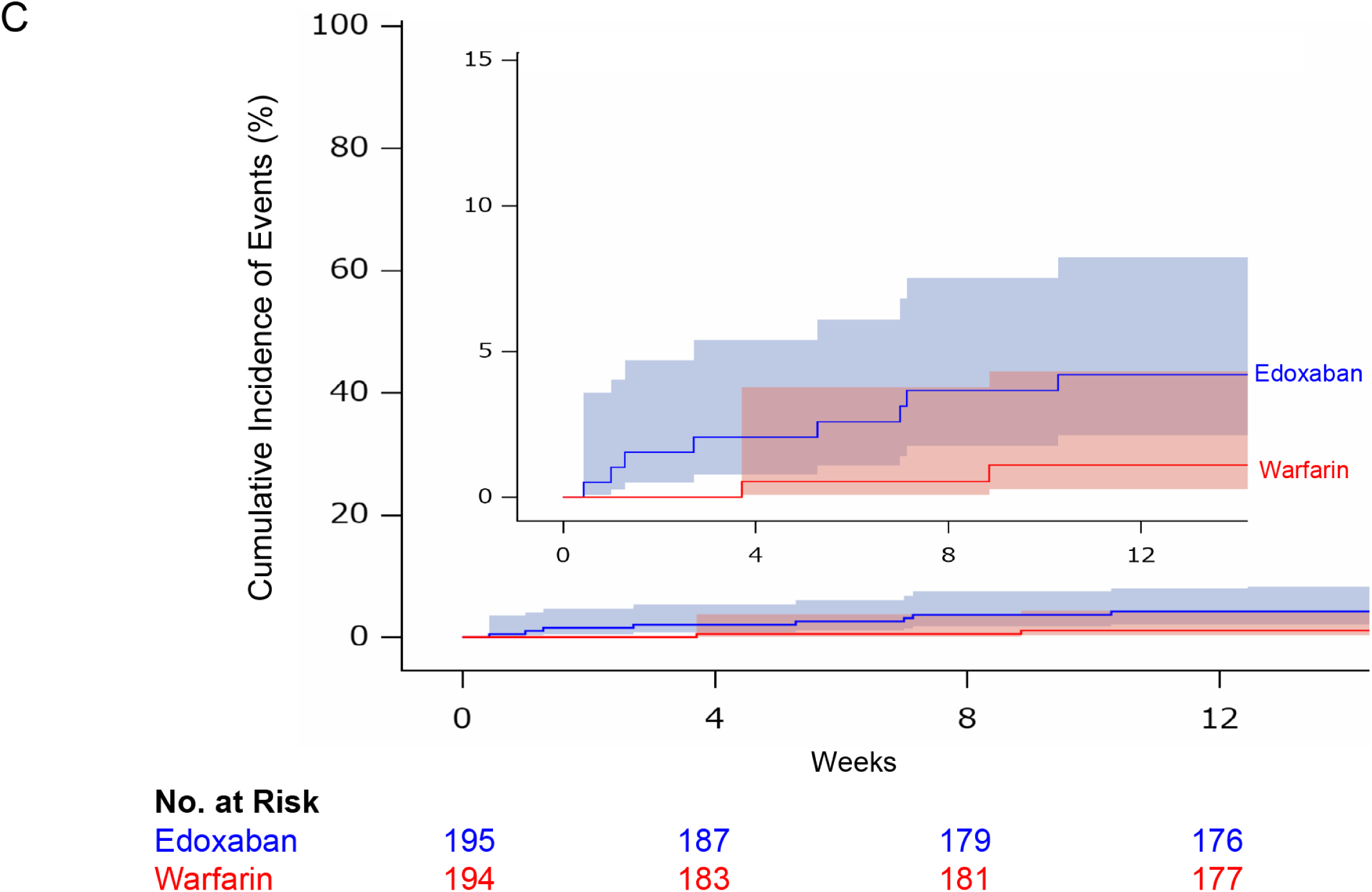
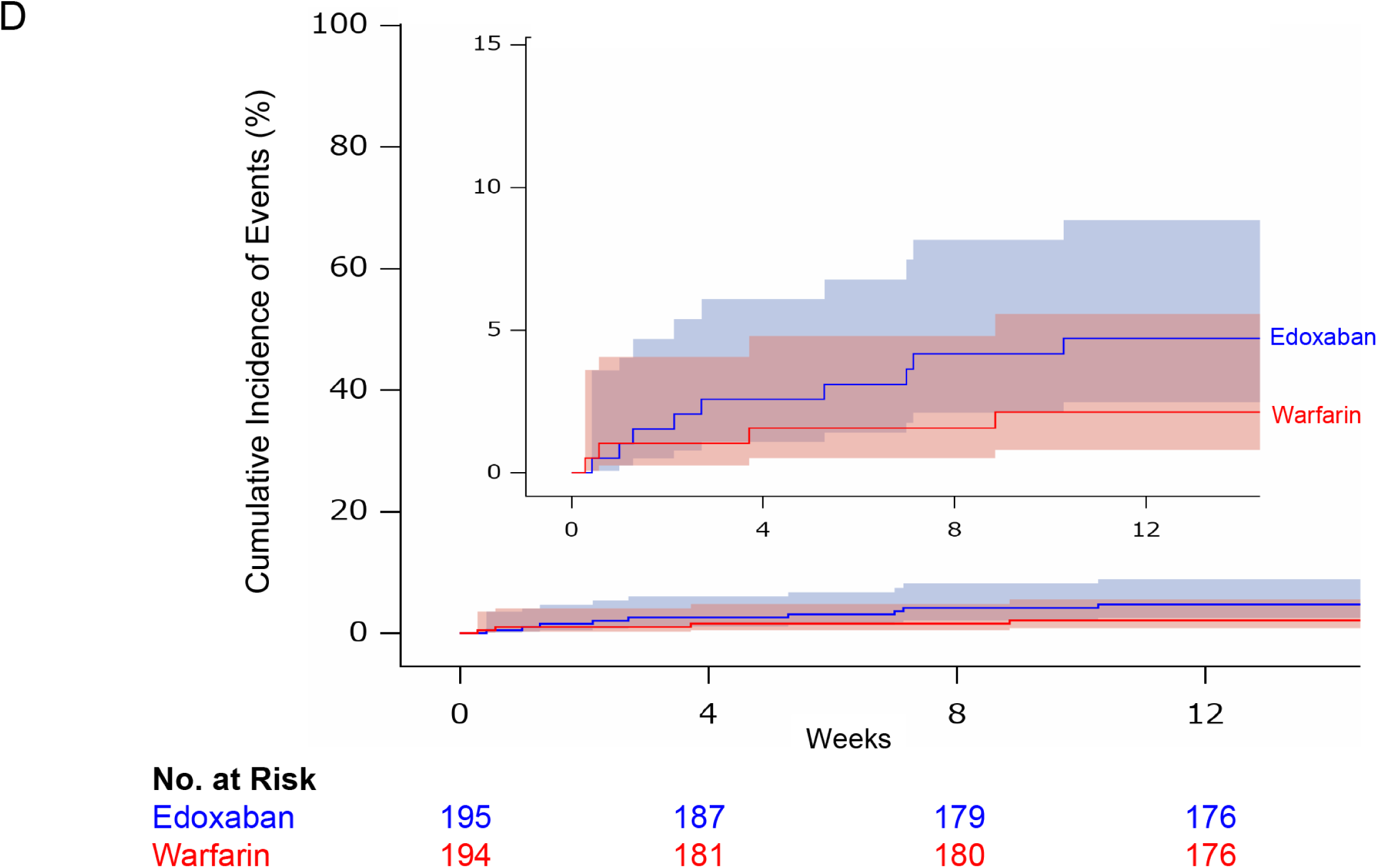
Kaplan–Meier analysis of the primary and key secondary endpoints. A: Primary endpoint. B: Intracardiac thrombus. C: Major bleeding. D: Net outcome (composite of stroke, systemic embolism, or major bleeding).

Major bleeding occurred in eight patients (4.1%) in the edoxaban group and in two patients (1.0%) in the warfarin group (risk difference, 3.07; 95% CI, −0.67 to 7.27) (Figures 2 and 3C). The sites of the major bleeding in each group are shown in Table S3. No fatal bleeding or intracranial hemorrhage was observed in patients treated with edoxaban despite high incidence of gastrointestinal bleeding. In the warfarin group, one patient died of cerebral hemorrhage. Clinically relevant hemorrhage occurred in 11 patients (5.6%) taking edoxaban and five (2.6%) taking warfarin (risk difference, 3.06; 95% CI, −1.46 to 7.84).

The net outcome (the composite of stroke, systemic embolism, or major bleeding) occurred in nine patients (4.6%) in the edoxaban group and in four patients (2.1%) in the warfarin group (risk difference, 2.55; 95% CI, −1.64 to 7.02) (Figures 2 and 3D).

The incidences of the other secondary outcomes were also similar for the two groups (Figure 2).

### Results of the subgroup analysis

The incidences of the primary endpoint in the two groups were generally consistent across all the prespecified subgroups (Figure 4). With respect to major bleeding and the net outcome, the results were also generally consistent across all the prespecified subgroups (Figures S1 and S2).

**Figure 4.**
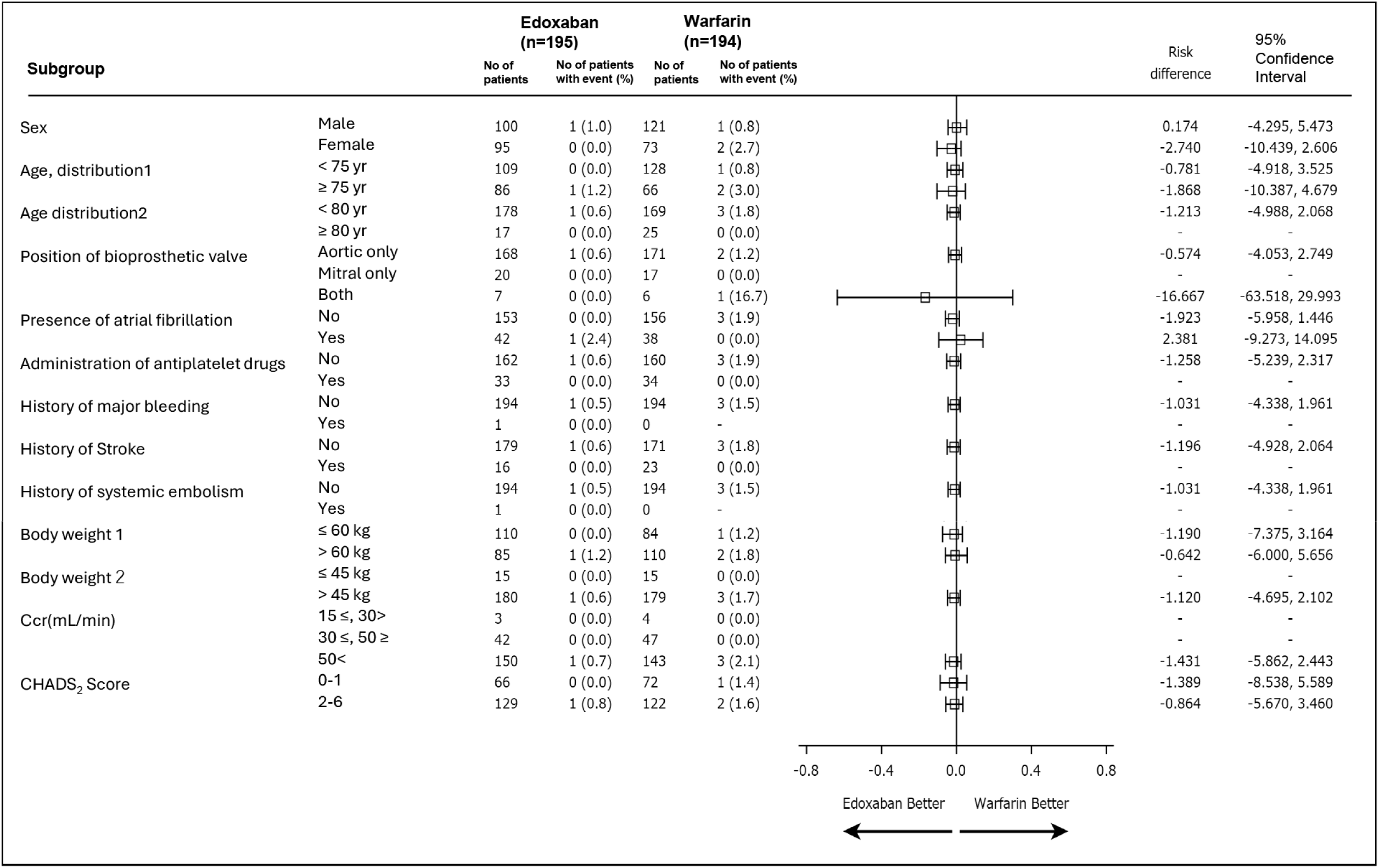
Results of the subgroup analysis of the primary endpoint Ccr, creatinine clearance.

## Discussion

The present study provides the first large-scale evidence on the use of DOAC in patients early after bioprosthetic valve surgery. Notably, approximately 80% of the study patients had sinus rhythm.

The incidence of embolic events has been reported to be high early after bioprosthetic valve replacement.^2–6, 22^ This high incidence of thromboembolic events may be caused by thrombus formation associated with the lack of endothelialization of prosthetic valves,^23^ the high prevalence of perioperative AF, and cardiac dysfunction early after bioprosthetic valve surgery. Therefore, current guidelines^7–9^ recommend anticoagulant therapy with vitamin K antagonists for 3 to 6 months following bioprosthetic valve replacement.

Warfarin has a narrow therapeutic range. Therefore, we need to adjust the dose of warfarin by blood monitoring of anticoagulation activity, and it takes a few days to enter the therapeutic range. In contrast, we can use constant dose of edoxaban determined by body weight, renal function and concurrent drugs, with no need of routine monitoring of anticoagulation activity. The effects of edoxaban emerge within 3 hours after administration, and edoxaban as well as other DOACs have a low risk of interaction with other drugs and food. The prevalence of DOAC administration has been increasing, and DOACs are prescribed in approximately 70% of patients with newly diagnosed AF, because of the aforementioned properties of DOAC.^24–26^ These properties are beneficial for patients early after cardiac surgery, when their condition is unstable. According to the results of a database analysis performed in the United States in 2020, the administration of DOACs at the time of discharge following bioprosthetic valve replacement had been increasing since 2011 in real-world clinical practice, despite this being off-label use in patients with sinus rhythm.^27^

Several studies have shown the efficacy and safety of DOACs in patients with a history of bioprosthetic valve replacement and AF.^13–19^ However, in the era of DOACs, there is a paucity of evidence regarding the efficacy and safety of DOACs in patients early after bioprosthetic valve replacement including patients with sinus rhythm.^20^

The results of the ENBALV trial demonstrate that edoxaban is comparable to warfarin with respect to the primary endpoint of stroke or systemic embolism in patients early after bioprosthetic valve replacement. The results of this clinical study met the primary endpoint, which was agreed by the Japanese Regulatory Pharmaceuticals and Medica Devices Agency. In addition, intracardiac thrombus did not occur in any of the patients taking edoxaban, but did occur in two patients taking warfarin. Thus, in patients early after bioprosthetic valve replacement, who are at a high risk of thromboembolic events, edoxaban showed efficacy compared to warfarin for the prevention of thromboembolic event and intracardiac thrombus formation.

The incidence of major bleeding events was numerically higher in the edoxaban group (4.1% vs 1.0%; risk difference, 3.07; 95% CI, −0.67 to 7.27). However, fatal bleeding or intracranial hemorrhage did not occur despite high incidence of gastrointestinal bleeding. On the other hand, one patient died of cerebral hemorrhage in the warfarin group. In addition, the incidence of major bleeding in the edoxaban group in the present study was similar to that identified in previous studies^2, 3, 5, 20, 28, 29^ evaluating clinical events early after bioprosthetic valve replacement.

The time in therapeutic range in the warfarin group was very short (19.0%) in the present study. During the unstable period immediately following open heart surgery, there is a significant risk of hemorrhage; therefore, surgeons tend to underdose patients, such that their PT-INRs are shorter than would be ideal. In addition, it is difficult to achieve appropriate therapeutic range of warfarin during the relatively short period of hospitalization. The short time in therapeutic range may influence on the occurrence rate of embolic events and bleeding. However, this undertreatment with warfarin reflects the current clinical situation and is precisely the problem associated with warfarin administration. Increase in treatment option of anticoagulant therapy early after cardiac surgery may have clinical advantages, because the conditions of the patients are highly variable during this period. It may give benefits for medical stuffs as well as patients, because edoxaban can be used with constant dose, no need of routine monitoring of anticoagulation activity, and a low risk of interaction with other drugs and food.

The present study had several limitations. First, the open-label protocol could have introduced bias. Warfarin should be administered with monitoring of dose adjustments using the PT-INR, whereas the dose of edoxaban is constant, determined by renal function and body weight, not the PT-INR. Therefore, we were unable to blind the participants or their physicians with regard to the treatment group. However, outcome assessments were conducted by assessors who were blinded to the treatment allocation. In addition, data management and monitoring were performed by independent clinical research entities to minimize the risk of bias. Second, the timing of the initiation of anticoagulant therapy was determined by the surgeons, which may have influenced the incidences of the clinical events. However, the present study was of patients in an unstable condition immediately after open-heart surgery, and therefore the study protocol was designed to prioritize patient safety. Third, we did not include patients undergoing transcatheter aortic valve replacement. However, the strategy of antithrombotic therapy after bioprosthetic valve replacement differs for patients who undergo surgical or transcatheter aortic valve replacement, according to the current guidelines.^7–9^ Therefore, the study population was limited to patients undergoing surgical bioprosthetic valve replacement.

In conclusion, edoxaban demonstrated comparable efficacy to warfarin for the prevention of thromboembolic events in patients early after undergoing bioprosthetic valve replacement, suggesting that it is a potential alternative anticoagulant therapy.

## Data Availability

The data collected for my study will not be made available to others except the investigators of this trial.

## Acknowledgments

We thank all the investigators (listed in the Supplemental Materials) and the study nurses/study coordinators involved in the ENBALV trial, and the staff members of DOT WORLD Co., Ltd for their assistance in the management of data collection, storage, and analysis.

We also thank Mark Cleasby, PhD from Edanz (https://jp.edanz.com/ac) for editing a draft of this manuscript.

## Supplemental material

List of investigators

Trial committee members

Tables S1–S3

Figures S1–S2

